## Supplementary figures and images for "A High Rate of COVID-19 Vaccine Hesitancy Among Arabs: Results of a Large-scale Survey"

### Supplementary Figure 1

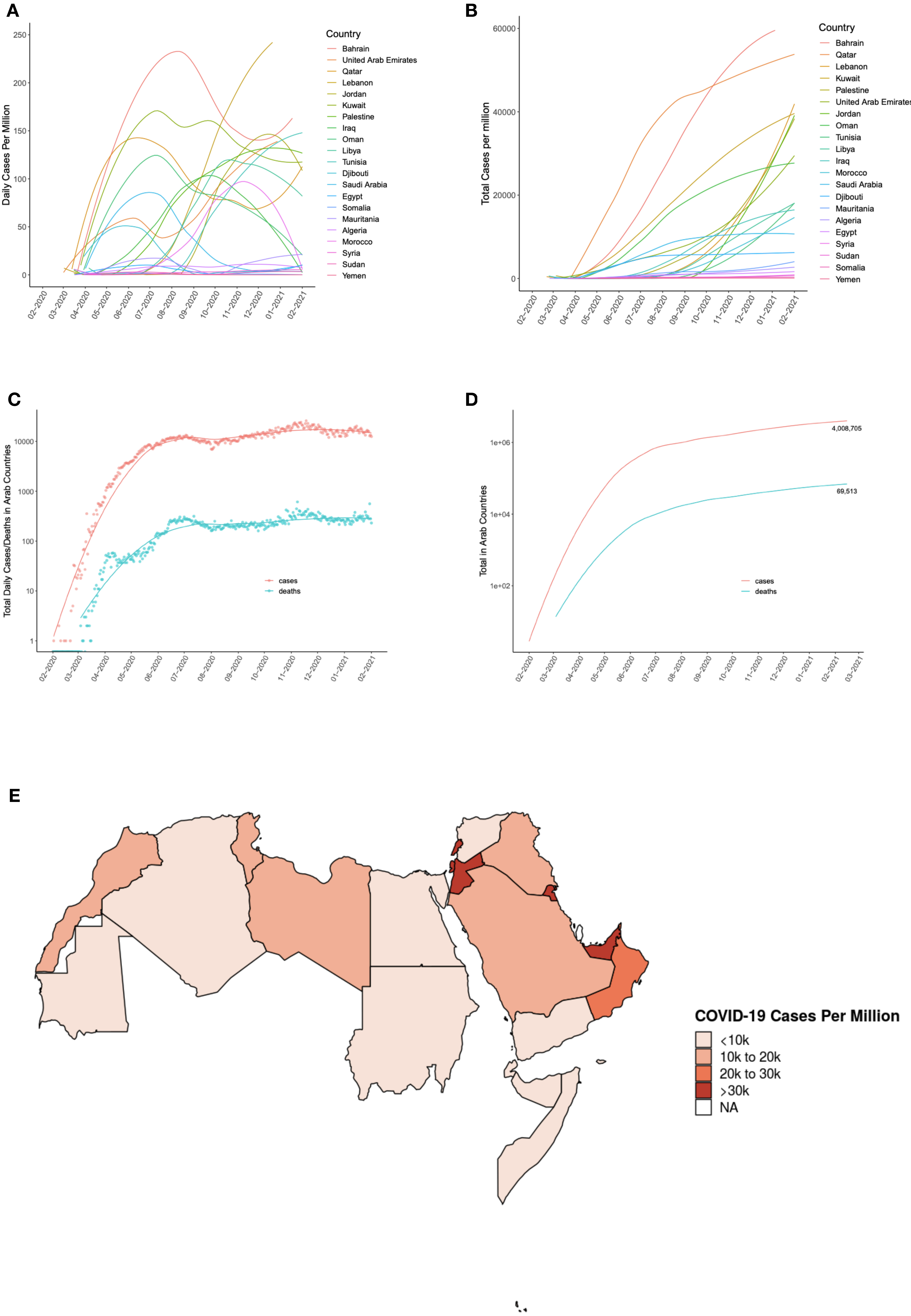

### Supplementary Figure 2

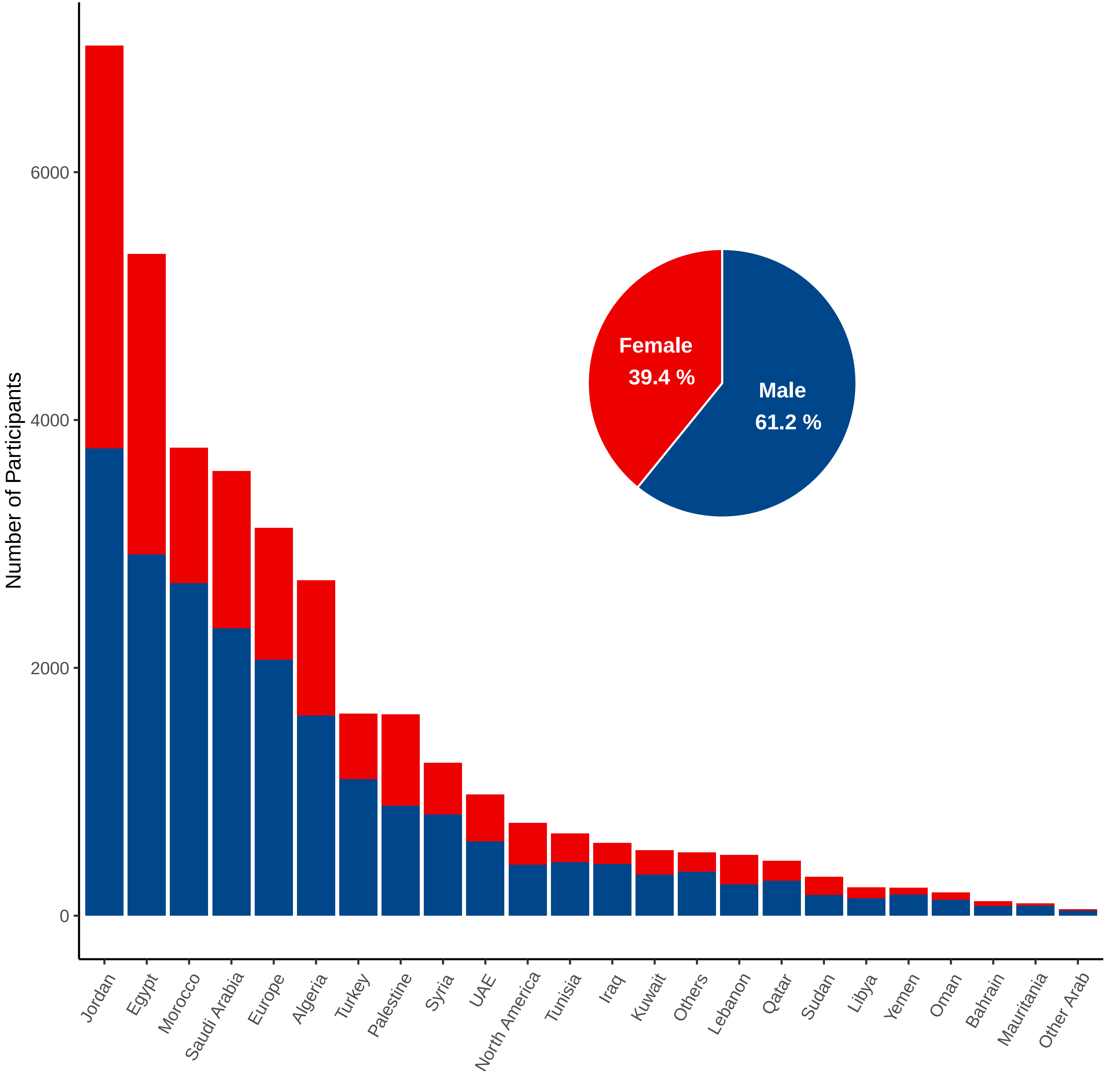

### Supplementary Figure 4

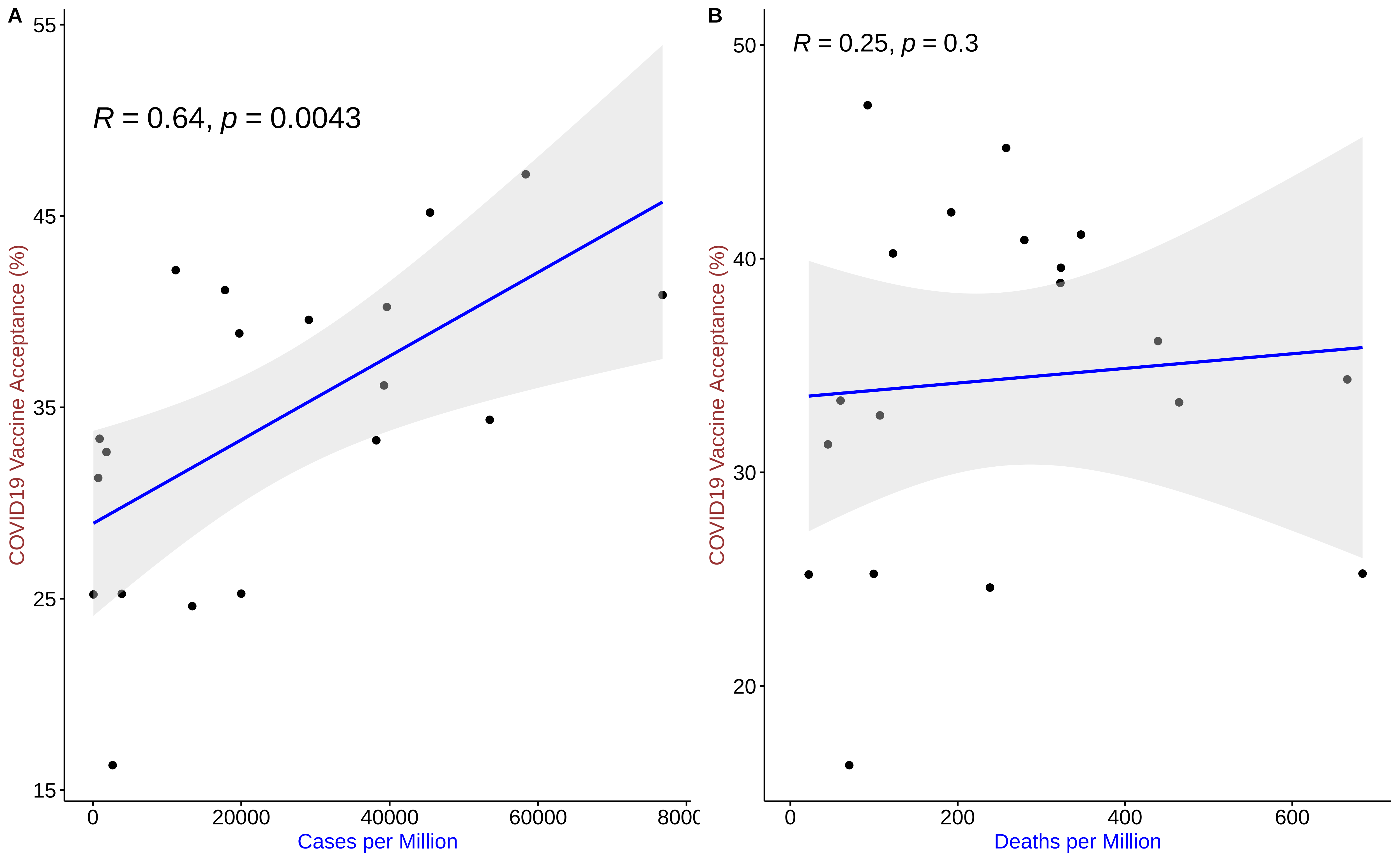

### Supplementary Figure 4

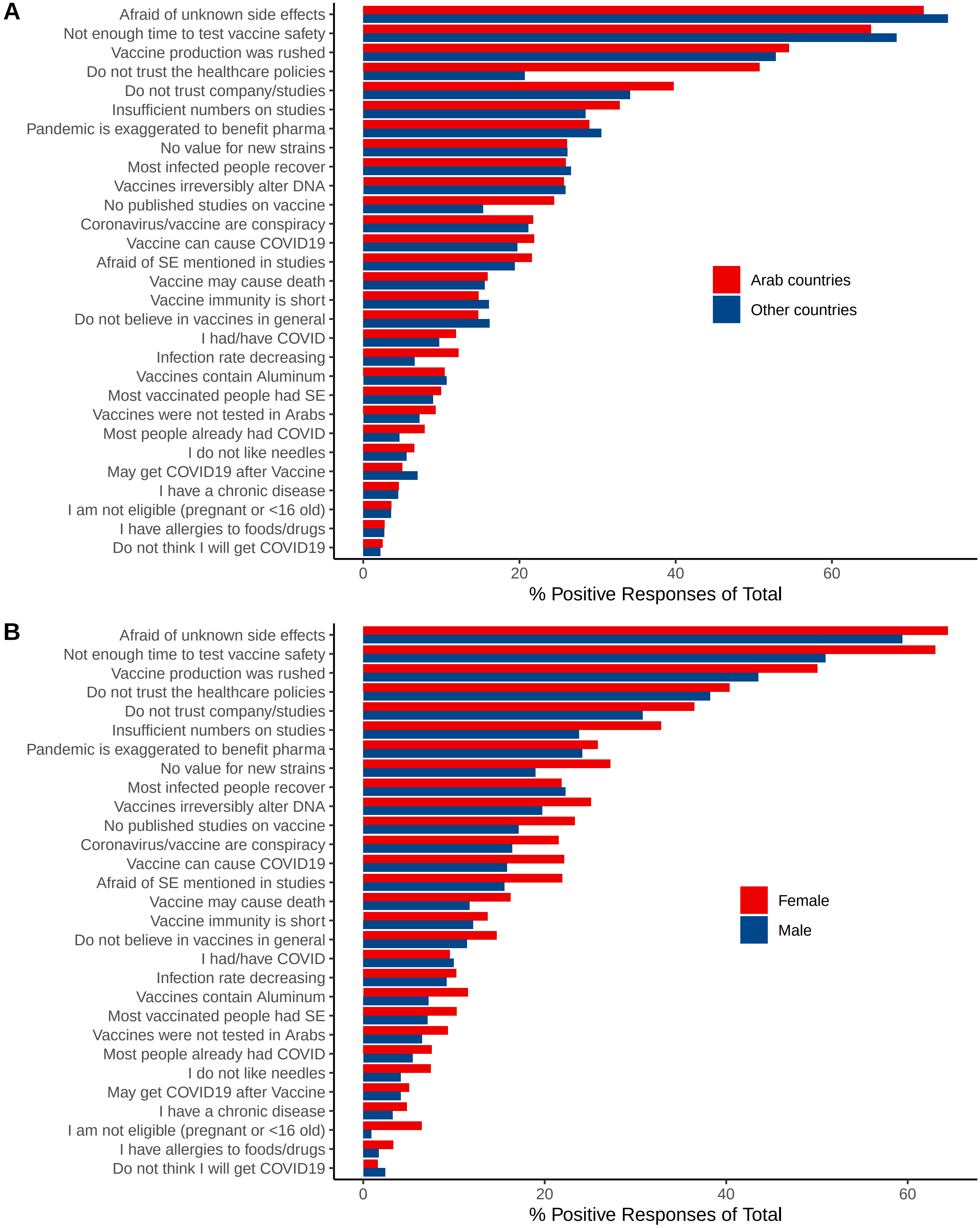
