## Supplementary Table 1 for "A High Rate of COVID-19 Vaccine Hesitancy Among Arabs: Results of a Large-scale Survey"

**Table 1 supplementary. Differences in Barriers for Acceptance according to Country of Residence, Gender and Academic Achievement, compared using chi-square test.**

| Question | Country | | | Gender | | | Academic Education | | |
|---|---|---|---|---|---|---|---|---|---|
| ` | Arab countries | Other countries | p | Female | Male | p | Higher education | Lower education | p |
| | 25884 (84.1) | 4895 (15.9) | | 12804 (41.6) | 17975 (58.4) | | 18942 (61.5) | 11837 (38.5) | |
| **I had/have COVID** | 2820 (86.7) | 434 (13.3) | NS | 1271 (39.1) | 1983 (60.9) | <0.0001 | 2124 (65.3) | 1130 (34.7) | <0.001 |
| **Afraid of unknown side effects** | 17505 (83.7) | 3405 (16.3) | <0.0001 | 8777 (42.0) | 12133 (58.0) | <0.0001 | 13211 (63.2) | 7699 (36.8) | <0.0001 |
| **I am not eligible (pregnant or <16 old)** | 841 (83.9) | 161 (16.1) | <0.0001 | 843 (84.1) | 159 (15.9) | <0.0001 | 591 (59.0) | 411 (41.0) | <0.0001 |
| **Infections numbers are decreasing** | 2971 (90.9) | 299 (9.1) | <0.001 | 1387 (42.4) | 1883 (57.6) | <0.0001 | 2104 (64.3) | 1166 (35.7) | <0.001 |
| **Vaccine production was rushed** | 13571 (84.6) | 2474 (15.4) | <0.0001 | 6899 (43.0) | 9146 (57.0) | <0.0001 | 10480 (65.3) | 5565 (34.7) | <0.0001 |
| **Most people already had COVID** | 1925 (90.0) | 215 (10.0) | <0.0001 | 1022 (47.8) | 1118 (52.2) | <0.0001 | 1301 (60.8) | 839 (39.2) | NS |
| **Most infected individuals recover** | 6395 (83.7) | 1242 (16.3) | NS | 2995 (39.2) | 4642 (60.8) | <0.0001 | 4746 (62.1) | 2891 (37.9) | <0.0001 |
| **Do not believe I will get COVID19** | 587 (86.3) | 93 (13.7) | <0.0001 | 204 (30.0) | 476 (70.0) | <0.0001 | 309 (45.4) | 371 (54.6) | <0.0001 |
| **Vaccines contain Aluminum** | 2579 (83.8) | 497 (16.2) | <0.0001 | 1583 (51.5) | 1493 (48.5) | NS | 1944 (63.2) | 1132 (36.8) | <0.0001 |
| **Do not believe in vaccines in general** | 3681 (83.0) | 756 (17.0) | <0.0001 | 2031 (45.8) | 2406 (54.2) | <0.05 | 2603 (58.7) | 1834 (41.3) | <0.0001 |
| **Coronavirus/vaccine are conspiracy** | 5463 (84.6) | 996 (15.4) | <0.0001 | 2977 (46.1) | 3482 (53.9) | <0.05 | 3851 (59.6) | 2608 (40.4) | <0.0001 |
| **No published studies done for vaccine** | 5991 (89.5) | 702 (10.5) | <0.0001 | 3156 (47.2) | 3537 (52.8) | NS | 4305 (64.3) | 2388 (35.7) | <0.05 |
| **Inuffcient numbers on studies** | 8083 (86.0) | 1312 (14.0) | <0.0001 | 4488 (47.8) | 4907 (52.2) | <0.05 | 6072 (64.6) | 3323 (35.4) | <0.001 |
| **Do not trust pharmaceutical/studies** | 9974 (86.0) | 1617 (14.0) | <0.0001 | 5060 (43.7) | 6531 (56.3) | <0.0001 | 7306 (63.0) | 4285 (37.0) | <0.05 |

| | | | | | | | | | |
|---|---|---|---|---|---|---|---|---|---|
| **Panedemic is exagerrated by pharma** | 7185 (83.5) | 1420 (16.5) | <0.001 | 3566 (41.4) | 5039 (58.6) | <0.0001 | 5614 (65.2) | 2991 (34.8) | <0.0001 |
| **No value for new strains** | 6231 (84.1) | 1176 (15.9) | <0.0001 | 3660 (49.4) | 3747 (50.6) | <0.0001 | 4759 (64.3) | 2648 (35.7) | NS |
| **may get COVID19 after Vaccine** | 1240 (79.6) | 318 (20.4) | <0.0001 | 697 (44.7) | 861 (55.3) | NS | 985 (63.2) | 573 (36.8) | <0.05 |
| **Vaccine immunity is short** | 3507 (83.4) | 700 (16.6) | <0.0001 | 1827 (43.4) | 2380 (56.6) | <0.0001 | 2895 (68.8) | 1312 (31.2) | <0.0001 |
| **Vaccine may cause death** | 3929 (84.4) | 725 (15.6) | <0.0001 | 2220 (47.7) | 2434 (52.3) | <0.0001 | 2825 (60.7) | 1829 (39.3) | <0.001 |
| **I have allergies to food/drugs** | 658 (84.5) | 121 (15.5) | <0.0001 | 435 (55.8) | 344 (44.2) | <0.05 | 502 (64.4) | 277 (35.6) | <0.0001 |
| **Most vaccinated people had SE** | 2415 (85.9) | 396 (14.1) | <0.0001 | 1390 (49.4) | 1421 (50.6) | <0.05 | 1679 (59.7) | 1132 (40.3) | NS |
| **Afraid of SE mentioned in studies** | 5276 (85.6) | 887 (14.4) | <0.0001 | 2965 (48.1) | 3198 (51.9) | <0.0001 | 3582 (58.1) | 2581 (41.9) | <0.0001 |
| **Vaccines irreversibly alter DNA** | 6388 (84.1) | 1209 (15.9) | <0.0001 | 3464 (45.6) | 4133 (54.4) | <0.05 | 4726 (62.2) | 2871 (37.8) | <0.0001 |
| **I have a chronic disease** | 1111 (84.4) | 206 (15.6) | <0.0001 | 653 (49.6) | 664 (50.4) | <0.05 | 833 (63.2) | 484 (36.8) | <0.0001 |
| **Vaccine can cause COVID19** | 5463 (85.5) | 926 (14.5) | <0.0001 | 3058 (47.9) | 3331 (52.1) | <0.0001 | 3703 (58.0) | 2686 (42.0) | <0.05 |
| **Afraid of needles** | 1517 (86.4) | 238 (13.6) | <0.0001 | 965 (55.0) | 790 (45.0) | <0.05 | 910 (51.9) | 845 (48.1) | <0.0001 |
| **Vaccines were not tested in Arabs** | 2255 (87.5) | 323 (12.5) | <0.0001 | 1264 (49.0) | 1314 (51.0) | <0.001 | 1578 (61.2) | 1000 (38.8) | <0.05 |
| **Not enough time to test vaccine** | 15887 (83.7) | 3102 (16.3) | <0.0001 | 8564 (45.1) | 10425 (54.9) | <0.05 | 12292 (64.7) | 6697 (35.3) | <0.0001 |
| **Do not trust health authorities** | 12373 (92.8) | 959 (7.2) | <0.0001 | 5512 (41.3) | 7820 (58.7) | <0.0001 | 8380 (62.9) | 4952 (37.1) | <0.0001 |
